# A First in Human Trial to Evaluate the Safety and Immunogenicity of a G Protein Based Recombinant Respiratory Syncytial Virus Vaccine in Healthy Adults 18-45 Years

**DOI:** 10.1101/2022.12.27.22283128

**Authors:** Xin Cheng, Gan Zhao, Aihua Dong, Zhonghuai He, Jiarong Wang, Brian Jiang, Bo Wang, Miaomiao Wang, Xuefen Huai, Shijie Zhang, Shuangshuang Feng, Hong Qin, Bin Wang

**Author notes:** Correspondence. Address:131 Dong An Road, 409 Fuxing Building, Shanghai 200032, People’s Republic of China.

## Abstract

**Background:** With enormous morbidity and mortality induced by respiratory syncytial virus (RSV) infection among infants and the elderly, vaccines against RSV infection are in huge market demand.

**Methods:** We conducted a First-in-human (FIH), randomized, double-blind, placebo-controlled dose escalation study to evaluate the safety and immunogenicity response of the rRSV vaccine (BARS13) in healthy adults aged 18-45. A total of 60 eligible participants were randomized in a 4:1 ratio to receive one of four dose levels or vaccination regimens of BARS13 or placebo.

**Results:** No serious adverse event (SAE) was experienced by any study participant. The geometric mean fold increase (GMFI) from baseline in all participants post-administration indicated that compared with single dose regimens, the IgG antibody level from baseline in participants received repeat dose regimens at Day 60 has increased over a 3-fold.

**Conclusions:** BARS13 has a generally good safety and tolerability profile, and no significant difference in terms of adverse reaction severity or frequency has been observed between different dose groups. The immune response in repeat dose recipients shows more potential in further study and has guiding significance for the dose selection of subsequent studies.

## Background

Respiratory syncytial virus (RSV) infection is a major cause of respiratory tract disease in children <5 years old. It leads to 64 million cases of bronchiolitis and viral pneumonia [1-5] and causes ∼200,000 deaths annually [4, 6]. A prophylactic treatment using palivizumab, and more recently, nirsevimab [7] can be used to prevent RSV in premature infants, infants with certain congenital heart defects or bronchopulmonary dysplasia, and infants with congenital malformations of the airway. However, antibodies’ economic cost limits its use to infants with identified risk factors residing in the developed world [8]. Besides the huge threat among the paediatric population, RSV infection is now recognized as a significant problem in elderly adults. Attack rates in nursing homes are approximately 5-10% per year, with substantial rates of pneumonia (10-20%) and death (2-5%). Estimates using US health care databases and viral surveillance results over a 9-year period indicate that RSV infection causes approximately 10,000 all-cause deaths annually among persons >64 years of age [9]. Although the vaccine is considered a more economical and effective strategy for preventing RSV-infected disease, no vaccine is currently available. This problem is left from the severity of the pathologic responses induced by vaccination with formalin-inactivated RSV (FI-RSV) to a large extent.

In the 1960s, the FI-RSV vaccine caused severe lung injuries in some paediatric recipients, and two infants died, resulting from a phenomenon that is now called vaccine-enhanced disease (VED) or enhanced respiratory disease (ERD) [10]. Pathological analysis showed that the dead infants had extended peribronchiolitis and alveolitis [11-13]. Subsequent studies have associated Formalin-inactivated RSV (FI-RSV) with low level of antibody responses and CD4+ T priming in the absence of cytotoxic T lymphocytes resulting in a pathogenic Th2 memory response with eosinophil and neutrophil [14], and such exacerbated T cell responses have been associated with immunopathogenesis of RSV in experimental models [15, 16]. Further understanding of the virus and VED mechanisms entails the new RSV vaccine designs toward being dependent on the induction of a robust, long-lasting neutralizing antibody response superior to the partial, transient immunity conferred by natural infection [17]. Various strategies to develop vaccines to target RSV have been tried. These include using a live-attenuated virus and various non-replicating virus components such as viral subunits and deoxyribonucleic acid (DNA) [18].

As of August 2022, global RSV vaccine development progress shows that only 1 Astra Zeneca RSV monoclonal antibody for paediatrics has been approved for marketing, 13 trials are at stages Phase II or Phase III, 11 trials are at Phase I, and more than a dozen candidates are still in preclinical phase [19]. More recently, one more AstraZeneca RSV antibody, nirsevimab, has been approved to prevent the RSV lower respiratory tract disease in newborns and infants on September, the 15^th^, 2022 [7]. The majority of phase II/III clinical trials are focused on paediatric and the elderly populations. In contrast, the adult population accounts for a minority, and only 3 trials are being carried out. The RSV vaccine based on F protein as the main target has been considered for RSV vaccine developments, including Pfizer and GlaxoSmithKline’s F protein-based RSV vaccine (for adults and the elderly) at phase III, while Janssen’s adenovirus vaccine and Merck’s RSV anti-F mAbs are also in the race.

Comparatively, the candidate vaccine (BARS13) is based on recombinant RSV viral G protein (RSV-G), containing two active components in an optimized ratio which are a purified RSV-G (expressed in E. coli system), which functions as the antigenic component, and cyclosporine A (CsA), which functions as an immunomodulator and the diluent to reconstitute the RSV-G. The G protein has been selected as the RSV immunogenic candidate as it has more stable neutralizing epitopes that are comparatively independent of its protein structure [20]. G protein functions as an attachment protein during RSV infection by interacting with the receptor of target cells. A monoclonal Ab against G protein has demonstrated activity inhibiting RSV infection in animal models [21]. CsA is a widely used immunosuppressant in organ transplantations and autoimmune diseases. It can induce antigen-specific T regulatory cells (Tregs) to ultimately achieve tolerogenic responses when combined with a protein antigen at a certain ratio and under a certain dose level [22]. In the development of BARS13, CsA was successfully used to generate tolerogenic responses with human PBMC in vitro [23]. As Treg plays an essential role in the suppression of VED [24], BARS13 was developed using a combination of the RSV-G with CsA to induce functional Tregs and a high level of neutralizing anti-RSV antibodies.

The preclinical studies have been performed in mice, rabbits, and rhesus macaque monkeys to investigate the immunological response to BARS13 and protective efficacy from the RSV challenge following immunization. It has been demonstrated that BARS13 not only induces a high level of neutralizing Abs against RSV, but also suppresses the exacerbated lung inflammation that occurs in animals vaccinated with either FI-RSV or G-protein based vaccines after a RSV challenge [25]. Based on these preclinical studies, we sought to test the safety, reactogenicity, and immunogenicity of the BARS13 investigational vaccine when administered intramuscularly (IM) to healthy adult participants aged 18 to 45 years.

## Methods

### Participants and study design

We conducted a phase I, first-in-human (FIH), randomized, double-blind, placebo-controlled dose-escalation study in healthy adults at a single center in Australia (Nucleus Network, Melbourne) from 16 October 2018 to 05 July 2019. The participants received a single or repeat vaccination schedule and different RSV-G protein plus CsA dose mixture. Participants, investigators, and laboratory staff were masked to treatment allocation. The primary objective was to evaluate the safety and reactogenicity of BARS13, and the secondary objective was to evaluate the humoral response in terms of immunoglobulin g (IgG) antibody levels to BARS13. The neutralization antibody response and T-cell response were evaluated as exploratory objective. The trial information can be obtained from Clinical Trial Registration (https://clinicaltrials.gov/ct2/show/NCT04851977).

Healthy males or females aged 18-45 years with no history of severe allergy or immunosuppressive therapy were screened for eligibility. All participants provided written informed consent before participation. Participants were enrolled and randomized in a 4:1 ratio sequentially using a dose-escalation protocol to receive BARS13 low dose (one injection of 9.2 μg rRSV-G protein/10 μg CsA to one arm, and one injection of saline/mannitol to the other arm), BARS13 high dose (one injection of 9.2 μg rRSV-G protein/10 μg CsA per arm) or placebo (one injection of saline/mannitol per arm). Both of the investigational vaccine and placebo have two vaccination regimens, single dose on Day 0 or repeat dose on Day 0 and 30 (Table 1). Among each cohort, 2 sentinels (n=1 active; n=1 placebo) will be assigned for a safety observation. In the absence of clinically significant safety signals in sentinel participants over a minimum period of 24 hours following vaccination, the remaining participants in the cohort could be vaccinated in a sequential manner. Enrolment into high-dose groups occurred only after a safety monitoring committee reviewed the data following vaccination of the participants in the previous low-dose group. Participants received vaccinations via intramuscular injection with RSV-G/CsA reconstituted solution or placebo according to a single (at day 0) or repeat (at days 0 and 30) vaccination schedule, with follow-up occurring for 60 days (all recipients) and 90 days (repeat dose recipients only) after the last vaccination.

**Table 1.** Study design.

| Step | Cohort | Treatment | Number of Participants | Vaccination Schedule |
| --- | --- | --- | --- | --- |
| I | 1 | BARS13 low dose single (LDS)<sup>a</sup> | 12 | Day 0 |
|  |  | Placebo<sup>b</sup> | 3 |  |
| II<sup>d</sup> | 2 | BARS13 low dose repeat (LDR)<sup>a</sup> | 12 | Days 0 and 30 |
|  |  | Placebo<sup>b</sup> | 3 |  |
|  | 3 | BARS13 high dose single (HDS)<sup>c</sup> | 12 | Day 0 |
|  |  | Placebo<sup>b</sup> | 3 |  |
| III<sup>d</sup> | 4 | BARS13 high dose repeat (HDR)<sup>c</sup> | 12 | Days 0 and 30 |
|  |  | Placebo<sup>b</sup> | 3 |  |

Note:
a. One dose of 9.2 µg rRSV-G protein/10 µg CsA by IM injection to the deltoid region of one arm, and one dose of placebo [saline/mannitol] by IM injection to the deltoid region of the other arm, given sequentially.
b. IM injection of saline/mannitol administered to the deltoid region of each arm (one injection of saline/mannitol per arm), given sequentially.
c. IM injection of 9.2 µg rRSV-G protein/10 µg CsA was administered to the deltoid region of each arm (one injection of 9.2 µg rRSV-G protein/10 µg CsA per arm), given sequentially. The high dose was twice the strength of the low dose.
d. If no safety holding criteria were met, initiation of both Cohort 2 and Cohort 3 (Step II; after review of Cohort 1 data) and Cohort 4 (Step III; after review of Cohort 3 data) could be triggered for enrolment at the discretion of the SMC.

### The vaccine

Advaccine Biopharmaceuticals Suzhou Co. Ltd in China manufactured the lyophilized powder of RSV-G and CsA diluent. The formulation buffer without active components was used as placebo. RSV-G lyophilized powder and vaccine diluent sterile solution are mixed together as the active BARS13 vaccine for injection. The information on study vaccine lots was listed in Additional file 1.

### Ethical compliance

This study was conducted in accordance to the principles of ICH-GCP, the Declaration of Helsinki, and applicable local regulations for conducting clinical trials on human medicinal products. This protocol was approved by the Alfred Hospital Ethics Committee. Human sera and PBMC were prepared in 360Biolabs in Malborne, Australia. Immunological tests were performed in Agilex Biolabs in Brisbane, Australia (anti-RSV G protein IgG antibody and neutralizing antibody) and Advaccine Biolabs in Suzhou, China (multiple cytokines assay and CD4 T cell proliferation test).

### Adverse events

The severity and relationship of adverse events (AEs) to the vaccine regimens were assessed by the investigators based on the US Food and Drug Administration (FDA) standards (*FDA 2007, Guidance for Industry: Toxicity Grading Scale for Health Adult and Adolescent Volunteers Enrolled in Preventive Vaccine Clinical Trials*). A placebo group was included in each cohort to serve as a comparative set that would facilitate the assessment of AEs potentially caused by the vaccine. Investigators were blinded to treatment assignment during the study to maintain unbiased assessment of AEs. The study participants were issued a daily diary card to capture treatment-emergent adverse events (arthralgia, diarrhea, fatigue, fever, headache, myalgia, injection site pain, swelling, and redness) during the 30-day follow-up period after each vaccination. Chemistry, haematology, and urinalysis were assessed using clinical samples (blood and urine) collected pre-vaccination on Days 0 and 30, and Days 7 and 30 after each vaccination. Vital signs were measured pre-vaccination at 30 and 60 minutes, and 7 days following each vaccination. Abnormal indicators of laboratory tests and vital signs were collected as AEs if accessed to be clinically significant.

The safety of BARS13 treatment regimens was based on the induction of adverse events (AEs) that represented both clinical and laboratory evaluations, using criteria that were pre-specified in the study protocol. We recorded treatment-emergent adverse events during the first 30 days as solicited TEAEs observation period after each vaccination, including the 30 minutes safety observation period post each vaccination and study days from 0 to 30 and from 30 to 60, and serious AEs (SAEs) until the last study visit.

### Safety data analysis

Since this is a pilot study, the sample size was determined based on practical and logistical considerations. A sample size of 60 participants is considered appropriate to achieve the defined objectives for the study. The safety population included all participants who received any study treatment (BARS13 or placebo) was used to perform safety and tolerability endpoints analysis. Basic descriptive analysis was used for each treatment regimen to present AEs. The per-protocol (PP) population consists of all participants in the immunogenicity population who received all vaccinations with BARS13 or placebo, without any major protocol deviations.

Within the PP set, the incidence of TEAEs, along with the ≥8% incidence rate of localized TEAEs and systematic TEAEs post each vaccination, were presented.

### Determination of anti-RSV G protein IgG antibodies with ELISA assay

Serum samples collected from all participants enrolled in the study on day 0 before vaccination and days 30 and 60 post-vaccination were applied for anti-RSV G protein IgG antibodies evaluation using a validated sandwich ELISA assay. Plates were coated with the rRSV protein G, followed by blocking. A standard RSV IgG serum (NIBSC, London, UK,Cat No.: 16/284) was serially diluted to set the standard curve ranging from 0.156 to 10.0 IU/mL. Human serum samples were diluted (at MRD of 1 in 1000) and added to the plate for 1hr incubation. After washes, goat anti-human IgG (H+L) peroxidase-labeled anti-protein G IgG antibodies (Invitrogen, Carlsbad, US, Cat No.: 31410) were subsequently applied to the plate for 1hr incubation and followed by washes. A colorimetric signal was developed by the addition of TMB (Sigma-Adlrich, St. Louis, US, Cat No.: T0440) and stop solution. The signal was read on an ELISA plate reader (SpectraMax VersaMax, Molecular Devices, Sunnyvale, US). The signal produced was proportional to the amount of analyte present and interpolated from the calibration curve presented on each plate. The concentrations of anti-RSV G protein IgG antibodies in samples were determined automatically by software SoftMax Pro (Molecular Devices, Sunnyvale, US, version 7.1) by reading off the calibration curves (4-PL curve fitting with 1/Y weighting factor). Data was then exported to Microsoft Excel (version 2021) and GraphPad Prism (GraphPad, San Diego, US, version 9.3) for further analysis.

All participants enrolled in the study were seropositive at baseline, showing a detectable level of anti-RSV-G IgG in their blood samples prior to the BARS13 administrated. Consequently, calculations of seropositivity and seroconversion rates were redundant. Due to this reason, the highest plasma dilution at which anti-RSV G protein antibodies were still detectable in the ELISA assay showed similar results at all assessed time points, including baseline (day 0 before vaccination) and days 30 and 60 per immunogenicity population in this study. Therefore, it was decided to evaluate the humoral response at a serum dilution equal to 1:2000.

### Neutralization antibody response with ELISA assay

The neutralization effect of the RSV Protein G vaccine on RSV infection was evaluated via a direct ELISA assay. The RSV envelope G glycoprotein contains a ∼40 amino acid central conserved domain (CCD; amino acid 162∼196) that lacks glycosylation and plays a critical role in virus infection and pathogenesis. RSV G CCD contains a CX3C motif that facilitates binding to the CX3CR1 receptor, leading to RSV infection in human airway epithelial cells. The previous study has shown that RSV G CCD is an exposed region that is accessible to antibody binding, and the antibody against this region could exhibit strain independence and neutralize RSV infection of human airway epithelial cells. Herein a direct ELISA assay was designed to evaluate the anti-CCD IgG level within the serum of participants, which could be considered as the surrogate of a traditional cell-based assay for neutralizing antibody evaluation. The assay format was similar to the Anti-RSV G Protein IgG Antibodies ELISA Assay. Plates were coated with RSV G CCD peptide amino acid 162∼196. A standard curve was normalized using NIBSC standard RSV IgG serum (Cat No.: 16/284) and ranged from 0.60 to 75.00 IU/mL. Human serum samples were diluted (at MRD of 1 in 200) and added to the plate. HRP Anti-Human IgG (Clone: G18-145) was applied as a detecting antibody. The plates were read at 450 nm and 620 nm on the VersaMax plate reader.

### CD4 T cell proliferation tested with flow cytometry method

Anticoagulant peripheral blood samples collected from all participants in cohort 2 and 4 on day 0 before vaccination, day 7 post first vaccination, and days 7 and 30 post the second vaccination, and lymphocytes were separated by Ficoll-plaque (Cityva, Logan, Utah, US) and cryo-freeze in liquid nitrogen for a long-term store. When lymphocyte was applied for CD4 T cell proliferation evaluation with a flow cytometry method, cells revived from liquid nitrogen tanks were assessed by live/death ratio and counted. Cells at 1×10^6^ for each well in a 96-well plate were cultured in cell incubator at 37°C with 5% CO_2_ for 120 hours and then stimulated by 100μL CD3/CD28 beads per well (Gibco, Grand Island, US, Cat No.: 11131D) as a positive control, 2 μg of RSV G peptide pools per well as antigen-specific stimulation, and 100μL M solution (RPMI1640 spiked with 40ng human-IL-2 (Peprotech, New Jersey, US, Cat No.: AF-200-02) and 40ng human-CD28 (MiltenyiBiotech, Bergesch Gladbach, Germany, Cat No.: 130-093-375)) as a negation control, respectively, for 120 hours in-vitro incubation in a cell incubator at 37°C with 5% CO_2_. These cells were stained with anti-human CD4-AF700 (Invitrogen, Cat No.: 5600488Z)/Fixable Viability Dye-eflour 780 (Invitrogen, Cat No.: 650865514) for 30 minutes, fixed and permeabilized with Fixation/Perm Diluent (Invitrogen, Cat No.: 00-5223-56/00-8333-56), intra-cellular staining with anti-human Ki67-BV421 (BD Bioscience, New Jersey, USA, Cat No.: 562899) for 1 hour, and then applied for data acquisition on Flow Cytometer (Attune NxT, Invitrogen, Carlsbad, US). Data was analyzed using FlowJo (BD Bioscience, version 10.6). Percentage of Ki67 positive cells gated from living CD4+ lymphocytes represented the proliferation of CD4 T cells.

### Multiple cytokines assay with beads based on flow cytometry method

Anticoagulant peripheral blood samples collected from all participants in cohort 2 and 4 on day 0 before vaccination, day 7 post first vaccination, and days 7 and 30 post the second vaccination were treated similarly as did in above T cell proliferation assay to generate the counted revived lymphocytes. Cells at 1×10^6^ per well in a 96-well plate were cultured in cell incubator at 37°C with 5% CO_2_ overnight (12 to 16 hours) and then stimulated by 100μL CD3/CD28 beads per well (Gibco, Cat No.: 11131D) as a positive control, 2 μg of RSV G peptide pools per well as antigen-specific stimulation respectively, for 24 hours in-vitro incubation in a cell incubator at 37°C with 5% CO_2_. Cell culture supernatants were collected and reacted with a commercial cytokine detection kit (Human Th Cytokine Panel with V-bottom Plate (Biolegend, California, US, Cat No.:741028)) for secreting cytokine (including IFN-γ, TNF-α, and IL-4) analysis. Data was acquired on Flow Cytometer (Attune NxT, Invitrogen) and analyzed using the LEGENDplex Software (Biolegend, version 8).

### Statistical analysis

No statistical hypothesis was formulated for this study and only descriptive statistics was performed for all parameters. Immunogenicity figures were plotted using GraphPad Prism 9. To delineate the GMC differences of binding antibody responses between each Cohorts, a Mann Whitney test was performed for ELISA results. A Kruskal Wallis test(H test)was performed for GMFI levels among Cohorts. All statistical tests were two-sided and differences with a *P* < 0.05 were considered significant.

## Results

### Study design

A total of 92 participants were screened for enrollment in this trial. Among them, 32 participants were excluded, and 60 eligible participants were enrolled and randomized. All participants received vaccination by BARS13 or placebo hence were included in the safety population. 56 (93.3%) were included in the immunogenicity population, and 53 (88.3%) were included in the per-protocol population (Fig.1).

**Figure 1.**
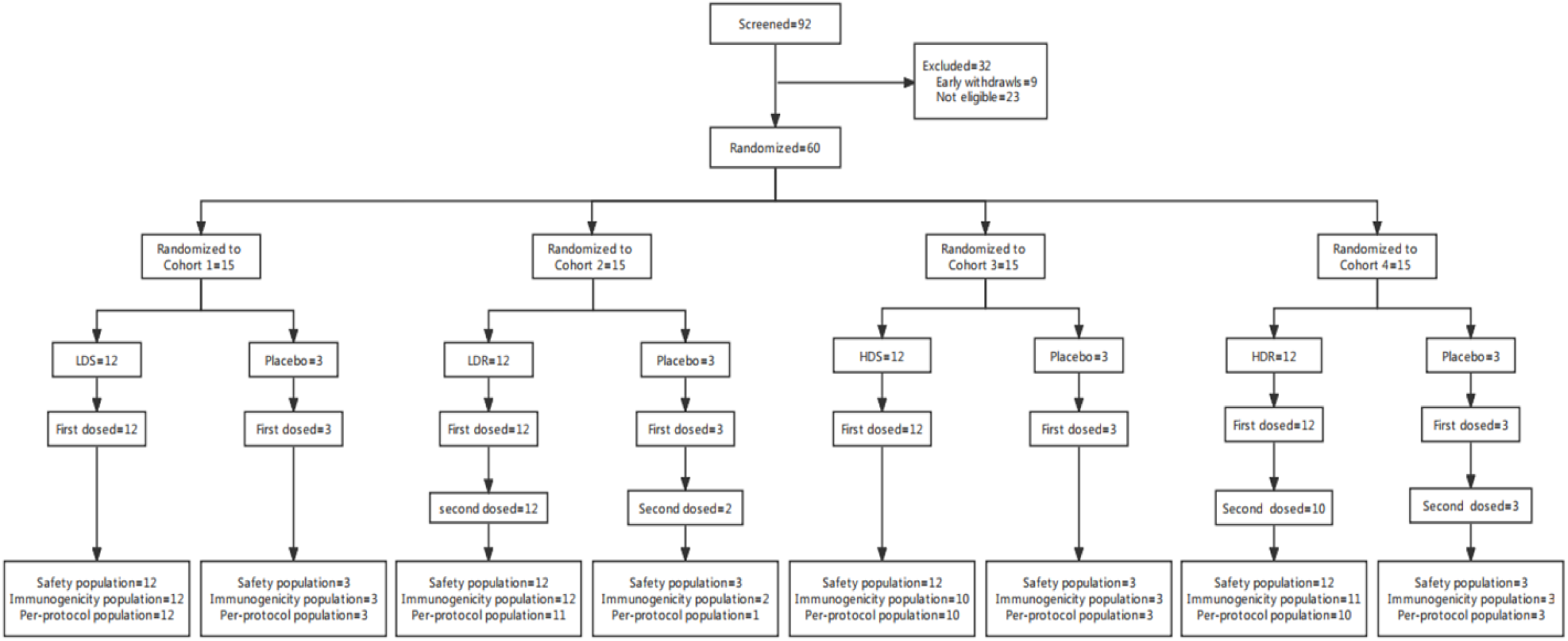
Study profile. Among each cohort, 2 sentinels (n=1 active; n=1 placebo) will be assigned for a safety observation at the study site for a minimum period of 60 minutes post-vaccination. Upon completion of the on-site safety assessments and release from the site, sentinel participants were monitored by follow up telephone call (at least one call) over a minimum period of 24 hours following vaccination. In the absence of clinically significant safety signals in sentinel participants over this period, the remaining participants in the cohort could be vaccinated in a sequential manner, with a minimum interval between participants of 30 minutes to allow monitoring of any acute events. During the vaccination period, the 7-day safety data of Cohort 1 will be reviewed by the safety review committee (SRC) if no safety concern has raised, the initiation of enrolment for both Cohort 2 and Cohort 3 (Step II; after review of Cohort 1 data) and Cohort 4 (Step III; after review of Cohort 3 data) could be triggered.1 placebo-receipt from Cohort 2 and 2 BARS13 recipients from Cohort 4 failed to complete the second vaccination on day 30, leading to exclusion in the per-population set.

The majority of participants were white females with 46 (76.7%) females, and 14 (23.3%) males. Demographics and baseline characteristics were comparable between participants vaccinated with BARS13 or placebo across all cohorts (Table 2).

**Table 2.** Demographics and Baseline Characteristics (All Participants Population)

| **Characteristics** | **LDS**<br>**( N=12 )** | **LDR**<br>**( N=12 )** | **HDS**<br>**(N=12)** | **HDR**<br>**( N=12 )** | **Pooled**<br>**Placebo**<br>**(N=12)** | **Overall**<br>**( N=60 )** |
| --- | --- | --- | --- | --- | --- | --- |
| Age (years) | 25.40 (3.99) | 27.30 (6.72) | 28.80 (7.16) | 27.60 (6.76) | 28.00 (2.22) | 27.40 (5.87) |
| BMI at screening (kg/m<sup>2</sup>) | 25.44 (4.86) | 24.11 (5.39) | 24.21(4.25) | 23.97 (3.21) | 24.65 (3.77) | 24.48 (4.73) |
| **Gender** |  |  |  |  |  |  |
| Female | 11 (91.7%) | 10 (83.3%) | 8 (66.7%) | 9 (75.0%) | 8(66.7%) | 46 (76.7%) |
| Male | 1 (8.3%) | 2 (16.7%) | 4 (33.3%) | 3 (25.0%) | 4 (33.3%) | 14 (23.3%) |
| **Ethnicity** |  |  |  |  |  |  |
| Hispanic or Latino | 1 (8.3%) | 2 (16.7%) | 1(8.3%) | 1 (8.3%) | 0 (0.0%) | 5 (8.3%) |
| Not Hispanic or Latino | 11 (91.7%) | 10 (83.3%) | 11 (91.7%) | 11 (91.7%) | 12 (100.0%) | 55 (91.7%) |

Notes: Data are mean (SD) or n (%).
Abbreviations: Low dose single (LDS), low dose repeat (LDR), high dose single (HDS), and high dose repeat (HDR).

### Vaccine safety

No SAE was experienced by any of the study participant at any time during the study. No TEAEs were classified as severe or life-threatening. No TEAE leading to study withdrawal during the 30-day follow up period after vaccination except 1 TEAE of moderate asthma exacerbation reported by a placebo participant in LDR. The majority of the TEAEs recorded were classified as mild. The frequency of TEAEs and drug-related TEAEs did not increase with vaccine dose level and frequency. The figures of the overview AEs post each vaccination were attached in the (Figure. S1, Figure. S2).

Local pain/tenderness was the most frequent solicited local adverse reaction in participants treated with active vaccine. Fatigue was the most frequently reported solicited systemic adverse reaction, and it was the most frequently reported as severe. The incidence rate of adverse reactions did not increase with vaccine dose level and frequency. Furthermore, the incidence rates of most local and systemic adverse reactions have shown a detectable decrease after the second vaccination at day 30 compared with those after the first vaccination at day 0, independently of the vaccine dose.

The majority of solicited adverse reactions were classified as mild and none were classified as life-threatening. After the first vaccination on day 0, the most frequent localized adverse reaction in 24 low dose recipients (LDS and LDR) and 24 high dose recipients (HDS and HDR) was localized pain/tenderness, with the incidence rate of 45.8% and 66.7%, respectively (Fig. 2a). 5 (20.8%) events of localized pain/tenderness were reported as moderate in high dose recipients. In 12 placebo-recipients, localized pain/tenderness (8.3%) was also reported after the first vaccination. Other localized adverse reactions were reported no more than 8.3% in all Cohorts. After the second vaccination on day 30, 3 localized adverse reactions were reported among 12 LDR recipients, 12 HDR recipients and 6 placebo recipients. 2 (16.7%) moderate localized pain/tenderness were reported in LDR and HDR recipients, respectively. 2 (16.7%) mild localized pain/tenderness were reported in LDR and HDR recipients, respectively. 1 (16.7%) mild localized pain/tenderness and 1 (16.7%) mild eccymosis/discoloration were reported in placebo recipients. While eccymosis/discoloration and swelling/induration in LDR and HDR recipients were reported no more than 8.3% after the second vaccination.

**Figure 2.**
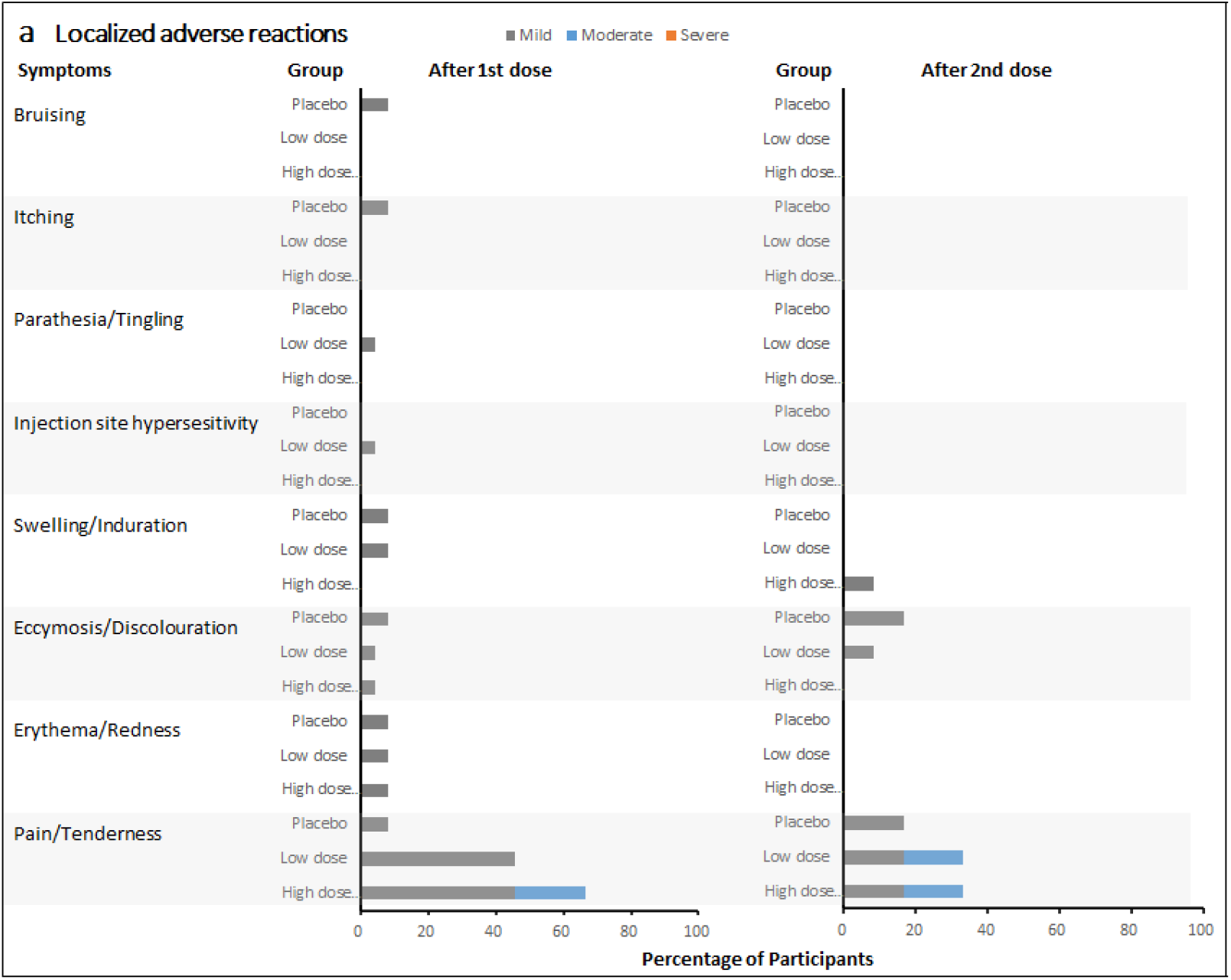
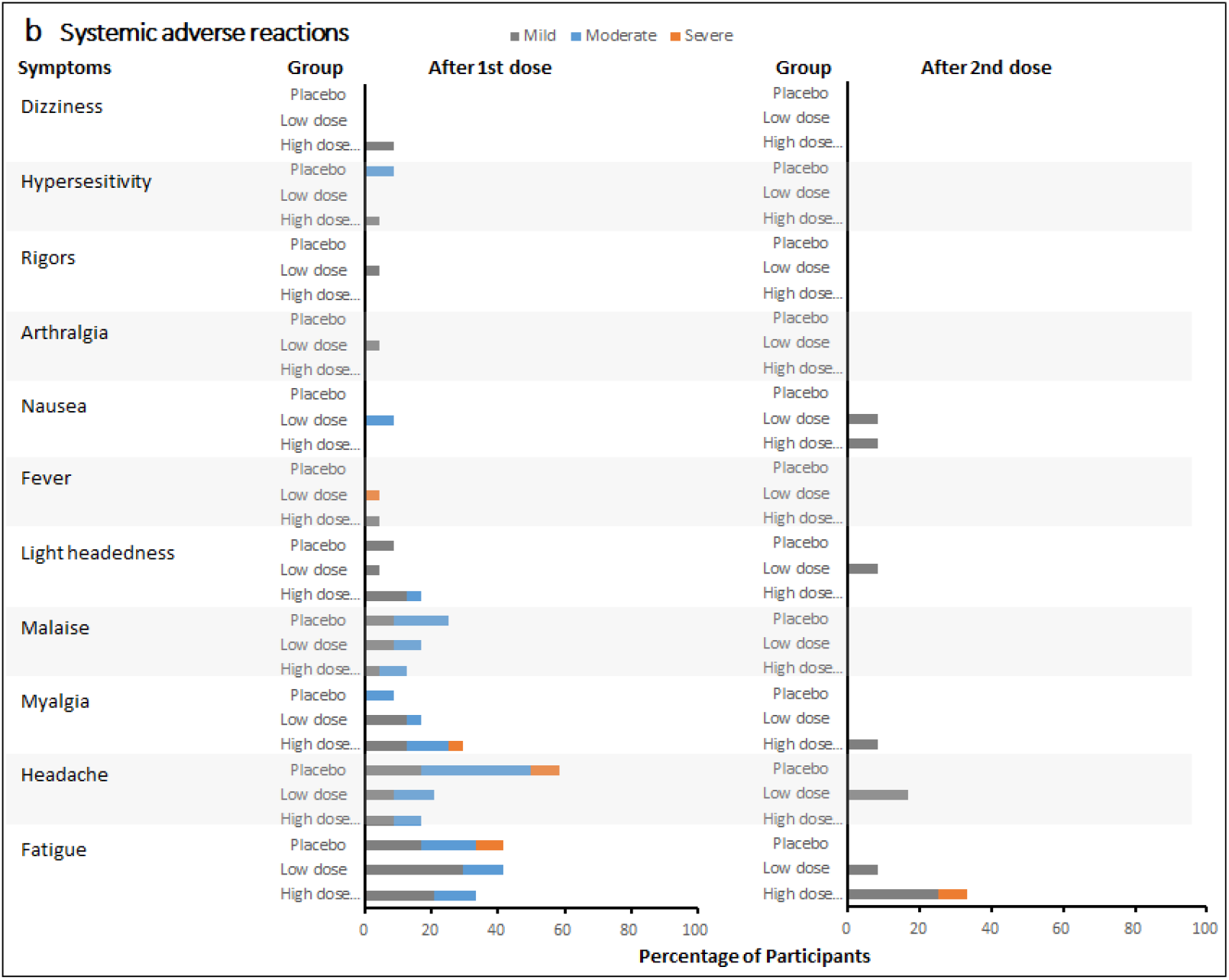
Incidence of localized and systematic adverse reactions in all Cohorts after each vaccination. a) Incidence of localized adverse reactions after each vaccination. b) Incidence of systematic adverse reactions after each vaccination.

After the first vaccination on day 0, the most frequent systematic adverse reactions were fatigue (41.7%), headache (20.8%), myalgia (16.7%) and malaise (16.7%) in low dose recipients (Fig. 2b). For high dose recipients, the most frequent systematic adverse reactions were fatigue (33.3%), myalgia (29.2%), headache (16.7%), light headedness (16.7%) and malaise (12.5%). The incidence of fatigue, headache and malaise were relatively high in the placebo recipients, with 5 (41.7%), 7 (58.3%) and 3 (25.0%) events reported respectively. Compared with all the mild and moderate fatigue and headache reported in both low and high dose recipients, 1 (8.3%) event of fatigue and 1 (8.3%) headache was each reported as severe in placebo recipients. No severe fatigue was reported in both low and high dose recipients. After the second vaccination on day 30, 1 (8.3%) fatigue was reported in HDR recipients. Other systematic adverse reactions were reported as mild in all Cohorts.

### Specific G protein binding antibody response

In the immunogenicity and per-protocol populations, the value of concentrations and the GMFI of G protein binding antibodies in terms of the BARS13 dosed cohorts on days 30 and 60 (repeat dose regimen only) were numerically higher with those on day 0 in anti-RSV-G IgG ELISA absorbance values using collected serum at 1:2000 dilution.

The median antibody concentrations at day 0, 30 and 60 in low dose recipients (Fig. 3a) and high dose recipients (Fig. 3b) each compared with placebo recipients were presented. The median antibody concentrations at day 30 was 1049.08 IU/ mL and 1126.61 IU/ mL for participants in LDS and HDS, respectively. The median antibody concentration of participants in LDR and HDR on day 30 was 763.18 IU/ mL and 885.74 IU/ mL, respectively. On day 60, antibody concentrations in the LDR and HDR were both higher than the baseline, with median values at 1187.10 IU/ mL and 1482.12 IU/ mL, respectively. From the distribution of antibody concentrations, after receiving 1 or 2 doses of BARS13, an apparent upward trend in the antibody concentration of all vaccine recipients was observed. Especially for participants who had a 2-dose-regimen (LDR and HDR), the increases of their IgG antibody concentrations were much higher at day 60 than themselves at day 30.

**Figure 3.**
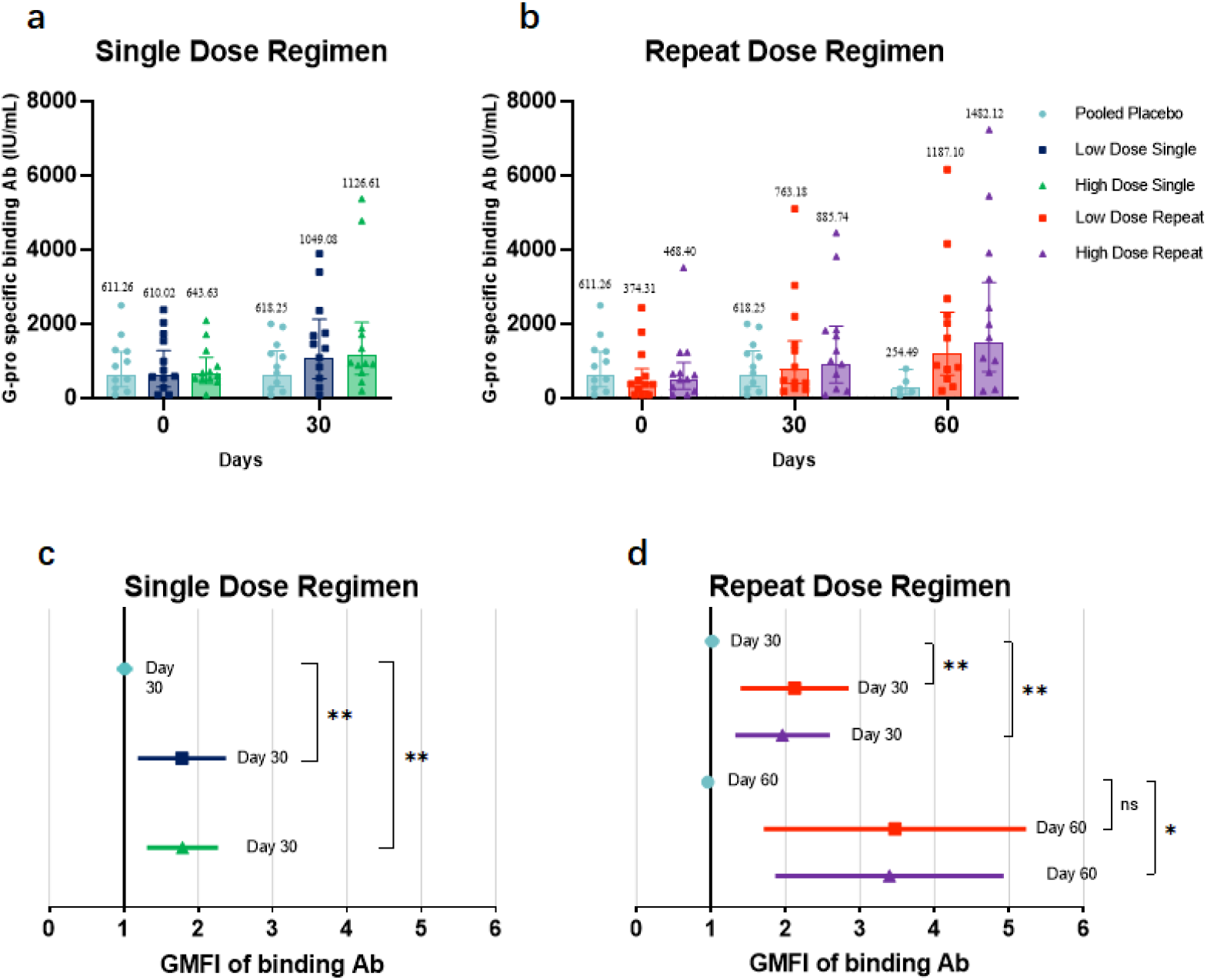
G-protein binding antibodies in all Cohorts. **a) The Geometric Mean Concentrations (GMCs) of binding antibodies in low dose recipients compared with in placebo recipients. b) The Geometric Mean Concentrations (GMCs) of binding antibodies in high dose recipients compared with in placebo recipients**. With ELISA test method, the concentrations of anti-RSV G protein antibodies in the plasma at days 0, 30, and 60 for high/low-dose and placebo cohorts were quantified using a standard curve prepared with the standard product antiserum to RSV (*P≤0.05, ns represents P>0.05). The original concentration is 2000 IU/ml. GMC and 95% confidence intervals are obtained from GraphPad Prism 9. **c) The geometric mean fold increase (GMFI) of binding antibodies in high dose recipients compared with in placebo recipients. d) The geometric mean fold increase (GMFI) of binding antibodies in high dose recipients compared with in placebo recipients**. The GMFI of BARS13 IgG antibody level in all cohorts at days 0, 30, and 60 (LDR and HDR only) and 95% confidence intervals are obtained from GraphPad Prism 9. P value is tested using Kruskal Wallis testing method (*P≤0.05, ns represents P>0.05).

The GMFI at day 0, 30 and 60 in low dose recipients (Fig. 3c) and high dose recipients (Fig. 3d) each compared with placebo recipients were presented. Descriptively, the GMFI for participants in LDS and HDS with BARS13 single dose at day 30 were 1.72 (95% CI: 1.23-2.41) and 1.75 (95% CI: 1.34-2.29), respectively. Both were higher than the placebo recipients with the GMFI of 1.01 (95%CI:0.92-1.11) at day 30. The GMFI for participants in LDR and HDR with BARS13 two-dose regimen at day 30 were 2.04 (95% CI: 1.44-2.88) and 1.89 (95% CI: 1.36-2.63), while at day 60 were 3.17 (95% CI: 1.88-5.36) and 3.16 (95% CI: 1.99-5.03), respectively. Comparatively, the GMFI for participants received placebo at day 60 were much lower with the increase-fold down to 0.96 (95% CI: 0.91-1.01).

The antibody data analysis demonstrates that levels of anti-G antibodies elicited in all BARS13 recipients were superior to that of the placebo recipients at all sampling time points. The GMFI of all BARS13 recipients have not shown any obvious increase at day 30, while LDR and HDR participants who received two-dose regimen have showed a detectable increase in the concentration of binding antibodies at day 60, suggesting that the two-dose regimen was more advantageous in terms of generating higher binding antibodies against RSV. In terms of dose selection, the high-dose cohorts (HDS and HDR) showed higher anti-G antibody concentrations on days 30 and 60 (LDR and HDR only) than the low-dose groups (LDS and LDR), respectively. Based on the analysis of the GMFI of BARS13 binding antibodies, the increase of binding antibodies was positively correlated with the increased doses and dosage of BARS13.

### Neutralization antibody response

Per immunogenicity population, the neutralization antibody concentrations at day 0, 30 and 60 in in low dose recipients (Fig. 4a) and high dose recipients (Fig. 4b) each compared with placebo recipients were presented. The increase of neutralization antibody concentrations in HDS and HDR were observed 30 days after their last vaccination. The GMC value of neutralization antibody in LDR and HDR at Day 30 was 1161.1 IU/mL (95% CI:599.6-2248.4 IU/mL) and 1507.9 IU/mL (95% CI:1.8997-1.3637 IU/mL), respectively, showing a detectable high increase compared with baseline GMC value. At Day 60, the GMC value of neutralization antibody maintained increasing trend and was up to 1683.6 IU/mL (95% CI:904.1-3135.4 IU/mL) and 2499.6 IU/mL (95% CI:1195-5228.3 IU/mL), respectively.

**Figure 4.**
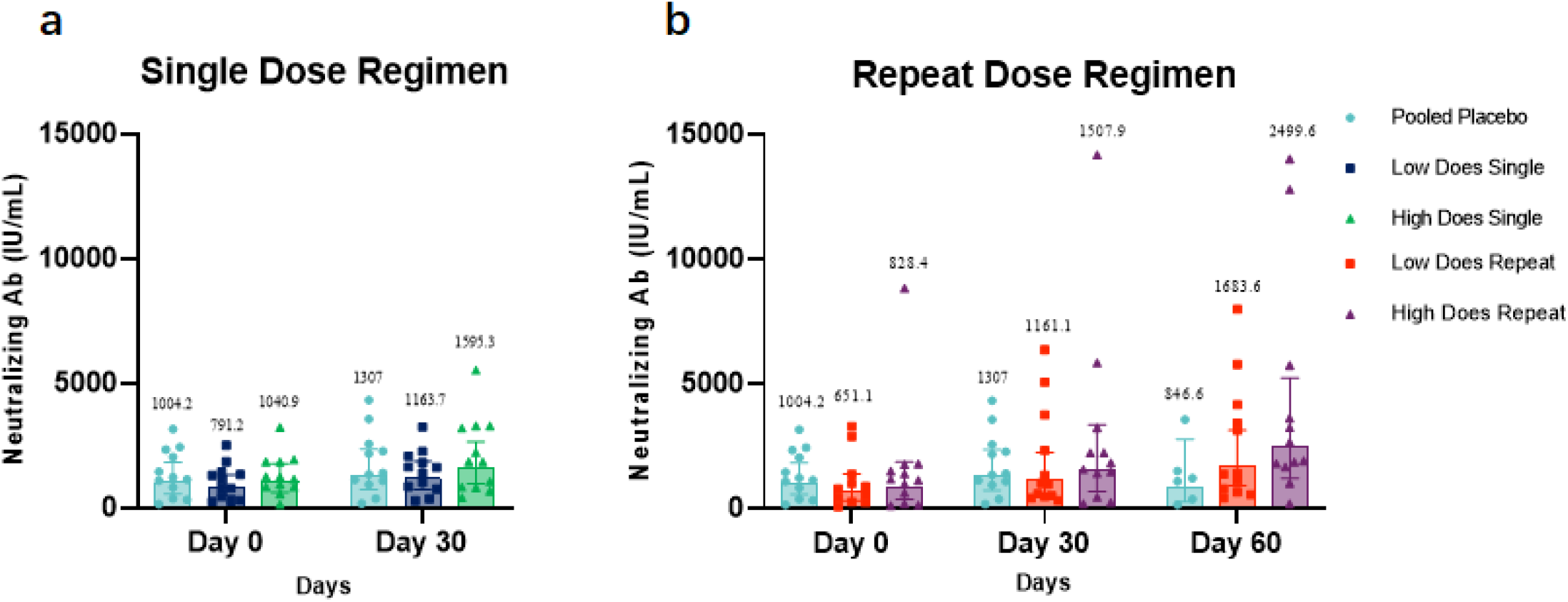
Neutralizing antibody tested with direct ELISA format in all Cohorts. **a) The Geometric Mean Concentrations (GMCs) of neutralization antibody in low dose recipients compared with in placebo recipients**. The GMCs of neutralization antibody of single dose regimens and placebo recipients at days 0, 30 and 60. Missing data were imputed using the Last Observation Carries Forward (LOCF) method. **b) The Geometric Mean Concentrations (GMCs) of neutralization antibody in high dose recipients compared with in placebo recipients**. The GMCs of neutralization antibody of repeat dose regimens and placebo recipients at days 0, 30 and 60.

The GMC value in LDS, HDS and participants dosed with placebo at Day 0 was 791.2 IU/mL (95% CI:476.6-1313.6 IU/mL), 1040.9 IU/mL (95% CI:618.9-1750.8 IU/mL) and 1004.2 IU/mL(95% CI : 552.7-1824.5 IU/mL), respectively. The GMC value with 95% CI of neutralization antibody in LDS, HDS and participants dosed with placebo at Day 30 was 1163.7 IU/mL (95% CI:725.1-1867.4 IU/mL), 1595.3 IU/mL (95% CI:964.7-2638.2 IU/mL) and 1307 IU/mL (95% CI:259.3-2764.3 IU/mL), respectively. The GMC value of neutralization antibody in participants dosed with placebo on days 0, 30, and 60 was 1004.2 IU/mL (95% CI:552.7-1824.5 IU/mL), 1307 IU/mL (95% CI:259.3-2764.3 IU/mL) and 846.6 IU/mL (95% CI:259.3-2764.3 IU/mL), respectively.

### Cellular Response

Having been demonstrated in animal studies, BARS-13 immunizations would induce Tregs that could suppress T cell proliferations when animals were exposed to RSV infection.^25^ To test if this is also true in human setting, we set up a flow cytometry method to explore proliferative profiles and functions of T-cells being restimulated in vitro by the RSV G peptide from LDR and HDR groups vaccinated by BARS13 in this trial. This test has been done in post hoc setting hence subjects have been unblinded and only subjects receiving BARS13 were included.

In LDR, compared with the high response readout stimulated by the CD3/CD28, as positive stimulants, the level of G-peptide stimulated IFN-γ and Ki67 were relatively stable with minimal increase after the second vaccination at day 37 and 60, while the level of G-peptide stimulated TNF-α has shown no obvious change from days 0 to 30 and actually has decreased at days 37 and 60. The level of IL-4 stimulated by the G-peptide was lower than that stimulated by CD3/CD28. The median and quartile values of IFN-γ stimulated by the G-peptide at days 0, 7, 30, 37 and 60 were 6.13 (2.33, 10.97), 1.93 (1.52, 3.35), 6.32 (6.32, 6.32), 1.78 (1.78, 1.78), and 1.25 (1.12, 3.99) pg/mL, respectively (Fig. 5a).

**Figure 5.**
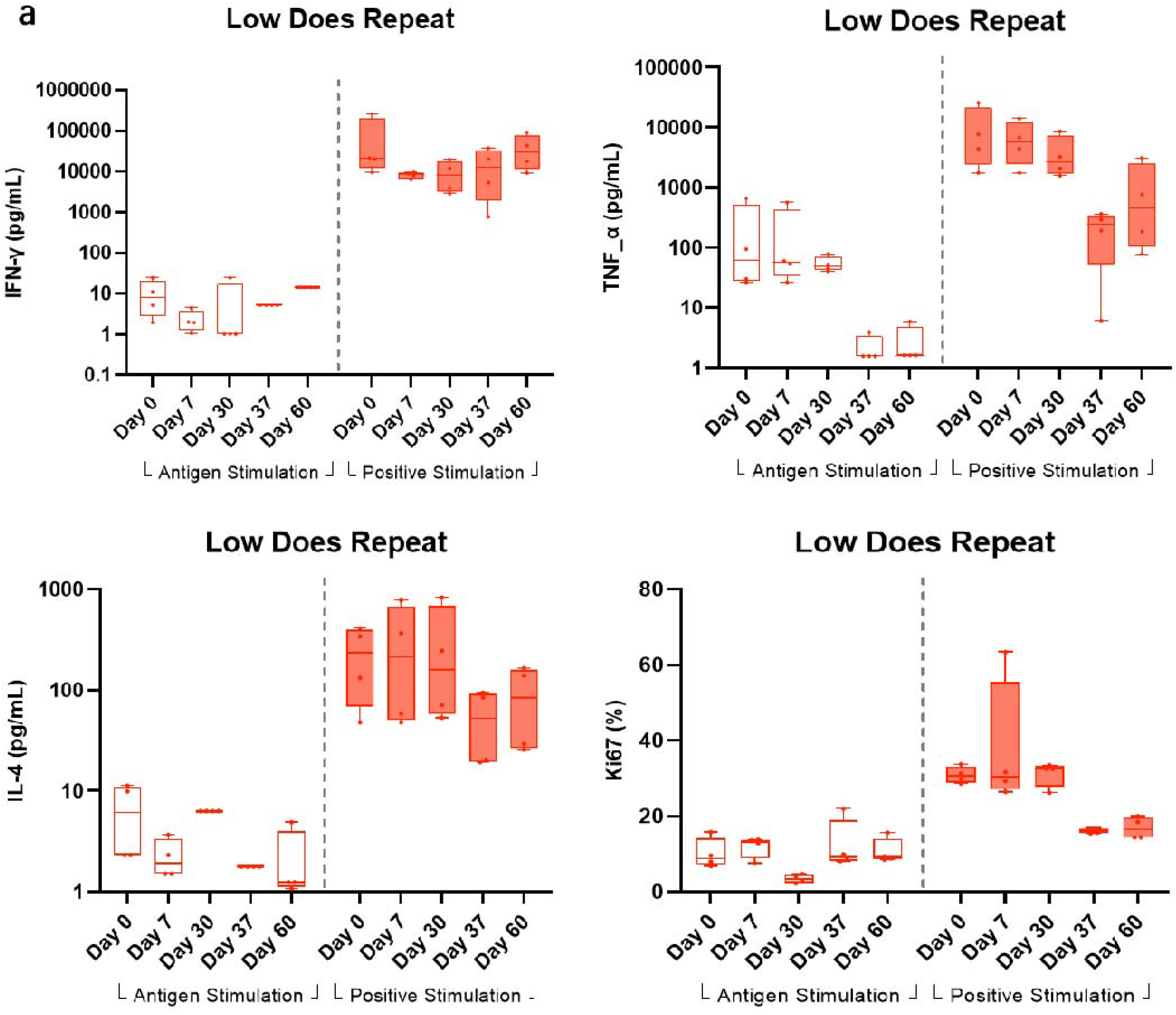
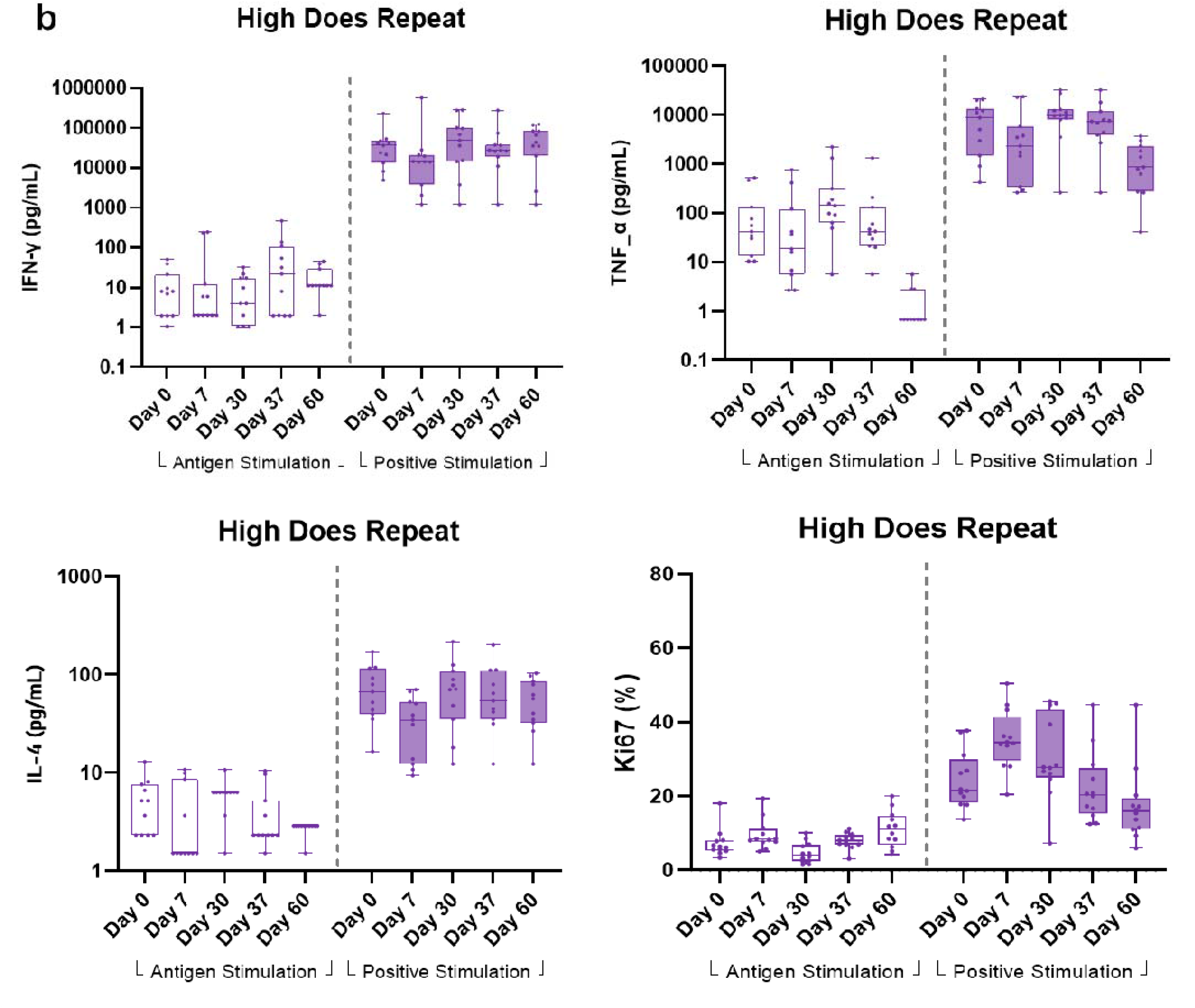
Specific T-cell response. a) T-cell response in low dose repeat recipients. The median and quartile values of Th1 cytokines (IFN-γ and TNF-α), Th2 cytokine (IL-4), and Ki67 in LDR were detected at days 0, 7, 30, 37 and 60 with the stimulation of G-peptide as specific antigen and CD3/CD28 as a positive control on cryopreserved PBMCs. Due to some samples on Day 0 were not recovered successfully resulting from long time (>1 year) cryopreservation, several subjects’ samples testing results lack baseline control and decision has been made that only ≥4 BARS13 recipients with their PBMC samples available at days 0, 7, 30, 37 and 60 in each Cohort were included in cellular response analysis. Therefore, 4 (33.4%) BARS13-reciptients in LDR has been included in the T-cell immunogenicity analysis. For the percentage readout of Ki67, a total of 11 (91.7%) BARS13 recipients in LDR were available for the analysis. **b) T-cell response in high dose repeat recipients**. The Th1 cytokines (IFN-γ and TNF-α), Th2 cytokine (IL-4), and Ki67 in HDR were detected at days 0, 7, 30, 37 and 60 with the stimulation of G-peptide as specific antigen and CD3/CD28 as a positive control on cryopreserved PBMCs. 4 (33.4%) BARS13-reciptients in HDR has been included in the T-cell immunogenicity analysis. All 12 BARS13 recipients in HDR were included in the Ki67 proliferation analysis.

In HDR, all the other Th1 related cellular cytokines (IFN-γ, TNF-α and Ki67) have shown no obvious change from pre-vaccination to post-vaccination timepoints for the G-peptide stimulated samples. Comparatively, CD3/CD28 stimulated samples have shown higher responses on the aforementioned cytokines. For Th2 biased cytokine IL-4, a consistent pattern with that of LDR is also shown, that positive stimulant samples also generate numerically higher readout across all timepoints (Fig. 5b).

## Discussion

We have performed a first-in-human phase I trial on BARS13, a novel designed RSV recombinant G protein vaccine with an immunomodulator, CsA. This RSV vaccine candidate was designed with the aim to suppress over-reactive T cells related to VED risk that has been observed in previous RSV vaccine clinical programs [25, 26]. Before this phase I trial, BARS13 vaccinations shown that RSV binding and neutralizing antibodies can be significantly increased and no VED symptom has been observed following detailed histopathology examination in a murine model RSV challenge study. This suppressed cellular response was associated with Tregs induction since Treg-knocked animals lost the ability to prevent VED in the same challenge study [25]. In a rabbit study, BARS13 vaccinations could induce long durable and recalled anti-RSV G antibody responses, but without T cells proliferations after being stimulated by the G peptide in vitro [26].

In this phase I trial, the majority of solicited local adverse reactions were classified as mild, and none as severe, nor life-threatening. No clinically significant vaccine-related safety or tolerability signals were reported during this study. The administration of BARS13 was generally tolerable, with no apparent differences between BARS13 and placebo vaccinated participants. The first-in-human study of the BARS13 has shown a tolerable and promising safety profile for this RSV vaccine candidate.

In the immunogenicity investigations, the anti-RSV-G IgG antibody titers measured by ELISA were expressed as concentration change from baseline and GMFI from baseline with 95% CIs for each of the individual treatment groups on days 30, and 60. For the absolute antibody levels as well as the fold increases from baseline to the post-vaccination, it is clearly exhibited a dose-dependent increase pattern from the low dose to high dose cohorts. Boost dose also contribute to the antibody response as shown in repeat dose cohorts, that after the second dose binding antibody has been increased further.

Previous studies have shown that vaccine-enhanced disease (VED) was due to over-reactive CD4 T cells, but it’s not clear how different CD4 T cell subsets lead to increased risk of VED [27]. In the meantime, human airway epithelial cells and alveolar macrophages can produce proinflammatory cytokines, such as tumor necrosis factor-α (TNF-α), which can help stimulate immune responses to inhibit RSV infection. Similar to children immunized with FI-RSV, BALB/C mice experimentally exhibited VED associated with Th2-biased immune responses [28]. In order to explore whether BARS13 vaccination will lead to proinflammatory cellular response, Th1-type cytokines (IFN-γ and TNF-α) and Th2-type cytokines (IL-4) were assessed by the flow cytometry to observe the T-cell immune response induced by BARS13 vaccination. From the PBMC testing results, it can be shown that the G-peptide stimulated PBMCs generally don’t show much obvious response to stimulation, but comparatively, anti-CD3/CD28 positive stimulant samples exhibited significant response to the in vitro stimulation, demonstrating that the responsiveness of PBMCs to external stimulant, and could potentially lead to the conclusion that BARS13 vaccination does not induce over-reactive T cell and had less chance to develop VED once RSV re-exposure occurs in those vaccinated subjects.

There are some limitations to this phase I study. First, limited follow-up period may not allow sufficient observation of potentially delayed reactions. Second, the methods for antibody testing, especially for neutralization antibody testing in Figure 4, further improvement such as benchmarking against international recognized standards which could make the results more comparable with other published RSV vaccine results is required. Third, due to extended long time cryopreservation of the collected PBMCs (more than 1 year), successful retrieval of cell samples was variable and rendered the result interpretations of flow analysis to be only tentative.

## CONCLUSION

In summary, the first-in-human trial has demonstrated that BARS13 not only induces meaningful level of anti-RSV-G Abs in a dose-dependent fashion, it also, importantly, demonstrates a well-tolerable and excellent safety profile from the recombinant RSV G protein with a low concentration of CsA in the formulation. This novel adjuvant, CsA, has provided potential of circumventing enhanced respiratory disease in RSV vaccine development history, which warrants further exploration in future clinical trials.

## Data Availability

All data produced in the present work are contained in the manuscript

## Supplementary Information

Additional file 1:

Figure S1. The incidence rate of TEAEs post 1^st^ vaccination by all cohorts

Figure S2. The incidence rate of TEAEs post 2^nd^ vaccination by repeat dose recipients

## Acknowledgements

We are grateful to Yongpeng Xu, Zhonghuai He, and team from Advaccine for their supports and contributions in the shipping of BARS13 and PBMC samples.

## Author Contributions

A.C., H.Q. and B.W., designed the clinical trial; A.D. and Z.H., developed manufacturing process, produced phase I clinical batch; S.Z. and S. F., participated in the development of flow cytometry assay; G.Z., J.W., B.J., B.W. and M.W., performed the ELISA and flow cytometry assays; A.C. and X.H., wrote the original draft; B.W., reviewed and edited the manuscript.

## Availability of data and materials

The data generated or analyzed during this study are included in this published article and its Additional files.

## Declarations

### Ethic approval and consent to participate

Not applicable. No private information of participants has been included in this article.

