## Supplementary 1 for "A First in Human Trial to Evaluate the Safety and Immunogenicity of a G Protein Based Recombinant Respiratory Syncytial Virus Vaccine in Healthy Adults 18-45 Years"

**Additional file 1.1 Study vaccine lots**

The following lot numbers of the study vaccines and diluents were used: Lyophilized rRSV-G protein 12µg (R201803); CsA diluent 20 µg/mL (C201802); Placebo for rRSV-G-protein (R201701K); Placebo for CsA (C201702K). BARS13 dose level was amended from 10 µg rRSV-G protein and 10 µg Cyclosporine A (CsA) to 9.2 µg rRSV-G protein and 10 µg CsA. During the dosing of 2 sentinel participants in Cohort 1, it was found that RSV-G was constituted in a lower volume than planned (0.42 rather than 0.5 mL) due to needle dead space was not accounted. By agreement between the Sponsor and the phase I site, it was decided to dose with 9.2 µg rRSV-G protein rather than 10 µg, while the total volume remained the same by adding extra diluent.
