## Supplementary Figure S1 for "A First in Human Trial to Evaluate the Safety and Immunogenicity of a G Protein Based Recombinant Respiratory Syncytial Virus Vaccine in Healthy Adults 18-45 Years"

**Additional file 2. Incidence rate of TEAEs post vaccination**

Incidence rate of TEAEs post 1st vaccination by all Cohorts

|  | LDS (N=12) | LDR (N=12) | HDS (N=12) | HDR (N=12) | Pooled placebo (N=12) |
| --- | --- | --- | --- | --- | --- |
| Any TEAE post 1st vaccination | 58.3% | 33.3% | 41.7% | 66.7% | 50% |
| Any TEAE during 30-minute post 1st vaccination | 16.7% | 8.3% | 8.3% | 8.3% | 41.7% |
| Any TEAE during 30-day follow-up period post 1st vaccination | 41.7% | 33.3% | 33.3% | 66.7% | 41.7% |
| Vaccine-related TEAE post 1st vaccination | 33.3% | 16.7% | 16.7% | 33.3% | 41.7% |

Abbreviations: Low dose single (LDS), low dose repeat (LDR), high dose single (HDS), and high dose repeat (HDR).

**Fig. S1** **The Incidence Rate of TEAEs post 1^st^ Vaccination by All Cohorts**
