## Supplementary Figure S2 for "A First in Human Trial to Evaluate the Safety and Immunogenicity of a G Protein Based Recombinant Respiratory Syncytial Virus Vaccine in Healthy Adults 18-45 Years"

**Additional file 2. Incidence rate of TEAEs post vaccination**

Incidence rate of TEAEs post 2nd vaccination by repeat dose recipients

|  | LDR (N=12) | HDR (N=12) | Pooled placebo (N=6) |
| --- | --- | --- | --- |
| Any TEAE post 2nd vaccination | 33.3% | 25% | 50% |
| Any TEAE during 30-minute post 2nd vaccination | 8.3% | 16.7% | 0% |
| Any TEAE during 30-day follow-up period post 2nd vaccination | 33.3% | 25% | 33.3% |
| Vaccine-related TEAE post 2nd vaccination | 16.7% | 16.7% | 0% |

**Fig. S2** **The Incidence Rate of TEAEs post 2^nd^ Vaccination by Repeat Dose Recipients**

Abbreviations: Low dose repeat (LDR) and high dose repeat (HDR).
